# A Bayesian approach for estimating typhoid fever incidence from large-scale facility-based passive surveillance data

**DOI:** 10.1101/2020.10.05.20206938

**Authors:** Maile T. Phillips, James E. Meiring, Merryn Voysey, Joshua L. Warren, Stephen Baker, Buddha Basnyat, John D. Clemens, Christiane Dolecek, Sarah J. Dunstan, Gordon Dougan, Melita A. Gordon, Robert S. Heyderman, Kathryn E. Holt, Firdausi Qadri, Andrew J. Pollard, Virginia E. Pitzer, the STRATAA Study Group

**Author notes:** **Corresponding author** Maile T. Phillips, P.O. Box 208034, 60 College St., New Haven, CT 06520-8034 USA.

## Abstract

Decisions about typhoid fever prevention and control are based on estimates of typhoid incidence and their uncertainty. Lack of specific clinical diagnostic criteria, poorly sensitive diagnostic tests, and scarcity of accurate and complete datasets contribute to difficulties in calculating age-specific population-level typhoid incidence. Using data from the Strategic Alliance across Africa & Asia (STRATAA) programme, we integrated demographic censuses, healthcare utilization surveys, facility-based surveillance, and serological surveillance from Malawi, Nepal, and Bangladesh to account for under-detection of cases. We developed a Bayesian approach that adjusts the count of reported blood-culture-positive cases for blood culture detection, blood culture collection, and healthcare seeking—and how these factors vary by age—while combining information from prior published studies. We validated the model using simulated data. The ratio of observed to adjusted incidence rates was 7.7 (95% credible interval (CrI): 6.0-12.4) in Malawi, 14.4 (95% CrI: 9.3-24.9) in Nepal, and 7.0 (95% CrI: 5.6-9.2) in Bangladesh. The probability of blood culture collection led to the largest adjustment in Malawi, while the probability of seeking healthcare contributed the most in Nepal and Bangladesh; adjustment factors varied by age. Adjusted incidence rates were within the seroincidence rate limits of typhoid infection. Estimates of blood-culture-confirmed typhoid fever without these adjustments results in considerable underestimation of the true incidence of typhoid fever. Our approach allows each phase of the reporting process to be synthesized to estimate the adjusted incidence of typhoid fever while correctly characterizing uncertainty, which can inform decision-making for typhoid prevention and control.

## Background

Current estimates of typhoid fever incidence serve as a basis for decision-making around typhoid control. However, facility-based cases of blood-culture-confirmed typhoid fever are considerably lower than the true number of those with the disease ^1^ because the reported numbers do not account for individuals with typhoid fever who do not seek healthcare, fail to receive a diagnostic test, or falsely test negative for typhoid (Fig 1). Annually, typhoid fever is estimated to cause 11-18 million infections and 100,000-200,000 deaths ^2-4^, but there is considerable uncertainty in these estimates.

**Figure 1.**
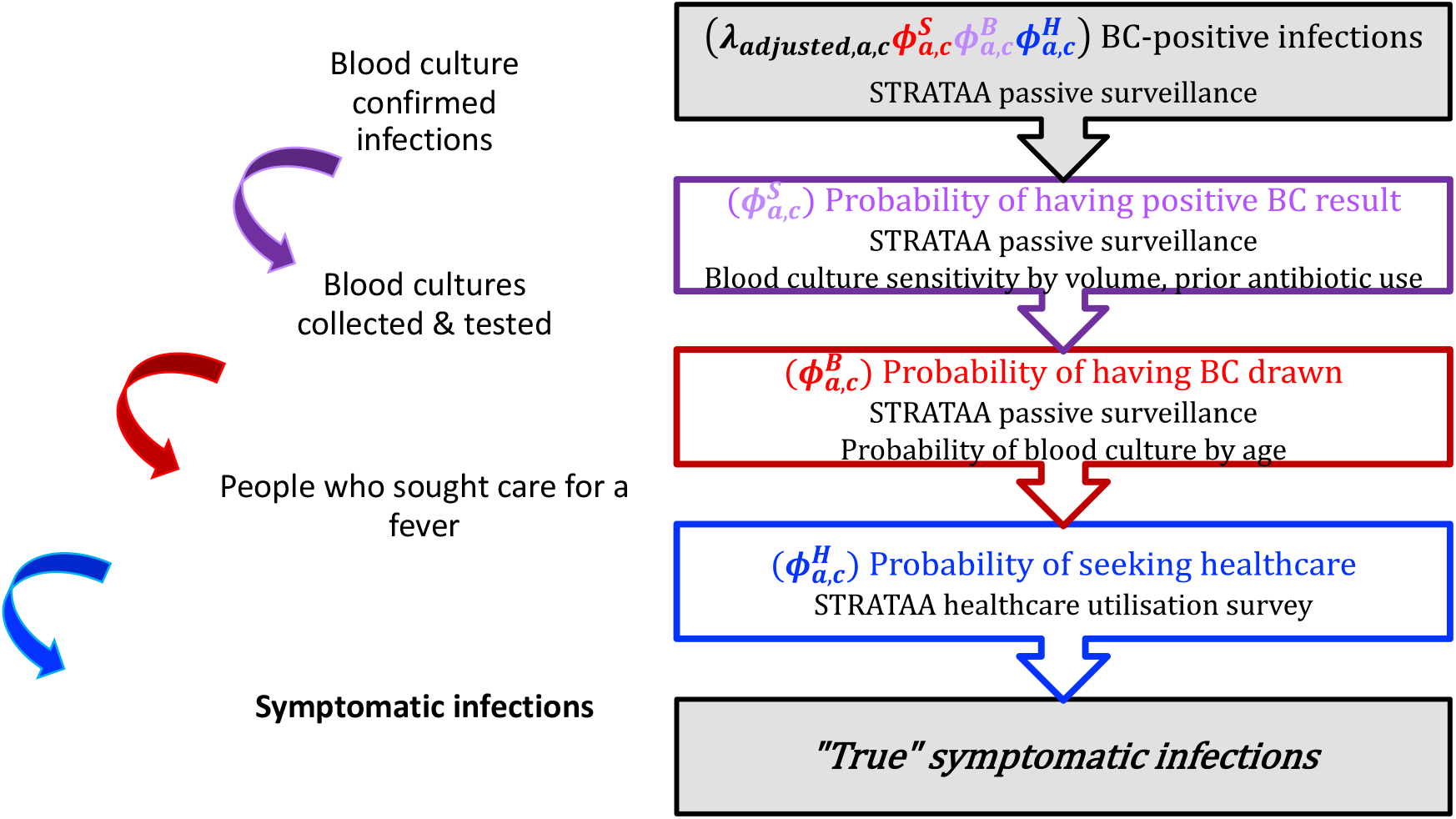
Flowchart of typhoid disease and observation process, and adjustment method to estimate the true number of cases. The pyramid (left) illustrates the different steps in the observation process for reporting typhoid incidence, with details on how parameters are estimated at each step. The flowchart (right) illustrates the corresponding Bayesian framework for each step of the observation process and which datasets and variables are used for adjustment. Adjustments for blood culture sensitivity are shown in purple, the probability of receiving a blood culture test is shown in red, and the probability of seeking healthcare is shown in blue. Variable definitions: *λ*, typhoid incidence rate; *ϕ*, a probability estimated in the model; *S*, sensitivity of blood culture; *B*, blood culture collection; *H*, healthcare seeking; *a*, age category; *c*, site. Abbreviations: BC, blood culture.

Studies suggest that somewhere between 60-90% individuals with typhoid fever do not receive adequate medical attention, in part because they do not to seek formal treatment ^5, 6^. Previous studies have found that healthcare utilization is correlated with the number of household members, distance to the healthcare facility, financial affordability, and trust in formal healthcare ^7, 8^. Furthermore, typhoid fever is often misdiagnosed based on physical examinations alone ^1^. Inconsistent clinical diagnoses arise because symptoms of typhoid, particularly prolonged fever, are also the main characteristics of other common infectious diseases in typhoid endemic settings ^1, 9^. Even if a blood culture test is recommended and laboratory facilities are available, not all patients will consent. Diagnostic tests can be invasive, and parents or guardians of young children sometimes do not want their children to have large amounts of blood drawn when they are already ill. In resource-poor countries in particular, lack of supplies and personnel lead to long wait times for receiving healthcare, further adding to lower rates of confirmatory testing. Clinical opinion on the cause of fever can also affect the likelihood of blood being drawn for culture ^10^.

Suboptimal diagnostic tests further contribute to underestimation of cases. Blood culture collection is the mainstay diagnostic test for typhoid fever ^11^, but it fails to capture approximately half of the true cases. The test sensitivity depends on the volume of blood drawn and whether a patient has recently received antibiotics ^12^. Thus, even if an individual with typhoid receives a blood culture test, he or she may falsely test negative and not be included in the reported number of confirmed cases.

The true incidence of typhoid fever cannot be directly assessed but can be estimated by accounting for steps in the reporting process. Methods to combine data from several sources to adjust for underestimation while accurately quantifying the uncertainty have been previously applied to estimate the incidence of HIV and influenza ^13-17^. Bayesian methods are conducive to integrating multiple data sources in this way. In this work, we developed a Bayesian multiplier framework to estimate population-based incidence of typhoid fever based on data collected from study sites in Africa and Asia.

## Methods

### Study design & data

We developed a framework within the Bayesian setting to integrate data from multiple sources to estimate the population-based incidence of typhoid fever based on passive surveillance in Malawi, Nepal, and Bangladesh, three typhoid-endemic countries with different demographics, healthcare systems, and access to diagnostics ^18^. Using this model, we sought to estimate the adjustment factors needed to calculate the “true” incidence of typhoid fever occurring in the population under surveillance, and to examine how these values varied by age across the three study sites, by combining information collected from the study population with estimates from prior published studies. We also compared our final estimates to serosurveillance data collected from the same population catchment areas.

Data came from the Strategic Typhoid Alliance across Africa & Asia (STRATAA) Programme, a prospective observational, population-based epidemiological study of typhoid incidence, transmission, and antibiotic resistance. From 2016-2018, the STRATAA investigators conducted demographic censuses, healthcare utilization surveys (HUS), passive surveillance, and serosurveys at each of three sites (Blantyre, Malawi; Kathmandu, Nepal; and Dhaka, Bangladesh). STRATAA’s study design and methods have been detailed elsewhere ^18^, and are briefly described below.

### Demographic census data

The demographic census was used to estimate the overall person-time contribution for incidence rate calculations. The survey documented household locations and individual characteristics for each geographically demarcated study area. The census provided information on each individual’s birthdate, sex, position in the household, marital status (if applicable), education level, and employment status (if applicable). Participants were surveyed and consented as households. Approximately 100,000 individuals were enrolled at each site, and census updates were carried out one to three times depending on the site. Over the two-year study period, this population amounted to 200,018 person-years of observation (pyo) in Malawi, 203,444 in Nepal, and 222,636 in Bangladesh.

### Passive surveillance

Clinical cases of culture-confirmed typhoid fever were identified through passive surveillance. Individuals living in the study areas who presented at partner facilities with a documented temperature of ≥ 38.0°C or a history of fever lasting at least two days upon presentation were eligible for enrollment. Healthcare workers collected clinical and demographic information from enrolled febrile individuals, as well as microbiological samples (urine, feces, and blood).

### Healthcare utilization survey

To estimate the proportion of cases captured by study facilities, we collected data from the head of household regarding actual and hypothetical usage of healthcare facilities for febrile episodes among household members from each of three age groups (<5 years, 5-14 years, >14 years), if available. Approximately 735 households were randomly chosen at each site, with the requirement that all households have at least one child (14 years or younger). The HUS contained questions regarding household and individual health behavior; house and household characteristics; water, sanitation and hygiene practices; and healthcare utilization (actual and hypothetical).

### Serosurveillance

Serosurveys were conducted in the census population to assess the underlying rate of seroconversion to typhoid, and to identify potential chronic carriers, initially based on anti-Vi immunoglobulin G (IgG). Approximately 8,500 participants from each site were randomly selected in an age-stratified manner. Healthcare workers collected serum samples from each individual upon enrollment and again three months later. Seroconversion was defined as a ≥ 2-fold rise in anti-Vi IgG titre between the first and second sample drawn and an absolute titre ≥50 EU/ml at the second time point to account for small variations above the lower limit of detection for the assay. We estimated the seroincidence by dividing the number of seroconversions by the person-time contribution between serum samples in each age group; 95% binomial confidence intervals (CI) were estimated.

### Approach and data analysis

We adjusted the reported number of blood-culture-positive cases of typhoid fever for each of the three phases of the reporting process: blood culture detection, blood culture collection, and healthcare seeking. We used information on healthcare seeking for fever and the proportion of fever cases who were enrolled and had blood taken for culture and combined this information with published data on risk factors for typhoid fever to reach our final adjusted numbers. We then examined whether our adjusted estimates were within the maximum range expected based on seroincidence estimates, which capture all clinical and sub-clinical cases.

The estimation of typhoid fever incidence is complicated by the relationship between fever and typhoid fever. In each phase of the reporting process, we can observe whether a person has a fever, but not necessarily whether he/she has typhoid fever. Thus, the symptomatic typhoid fever pyramid is nested within a larger fever pyramid (Fig S1).

We assumed that the observed number of blood-culture-positive individuals (*n*_observed,*a,c*_) follows a Poisson distribution given as

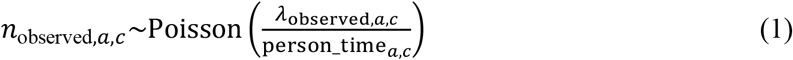

where *λ*_*observed,a,c*_ is the observed incidence rate of typhoid fever, adjusted for pyo from the demographic census (person_time_*a,c*_) (Fig 1, Table 1). Subscript *a* represents age category, where

**Table 1.**
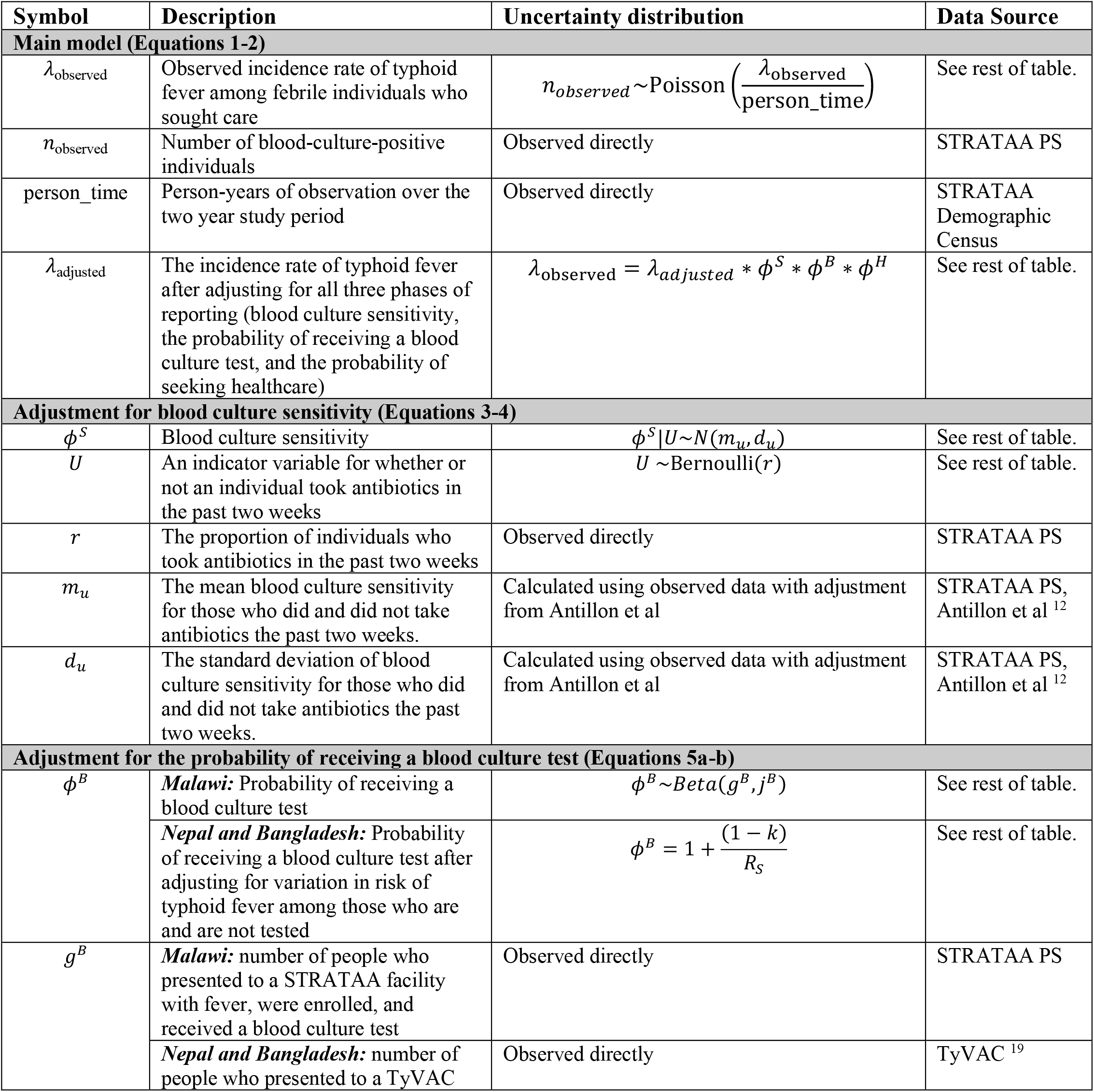
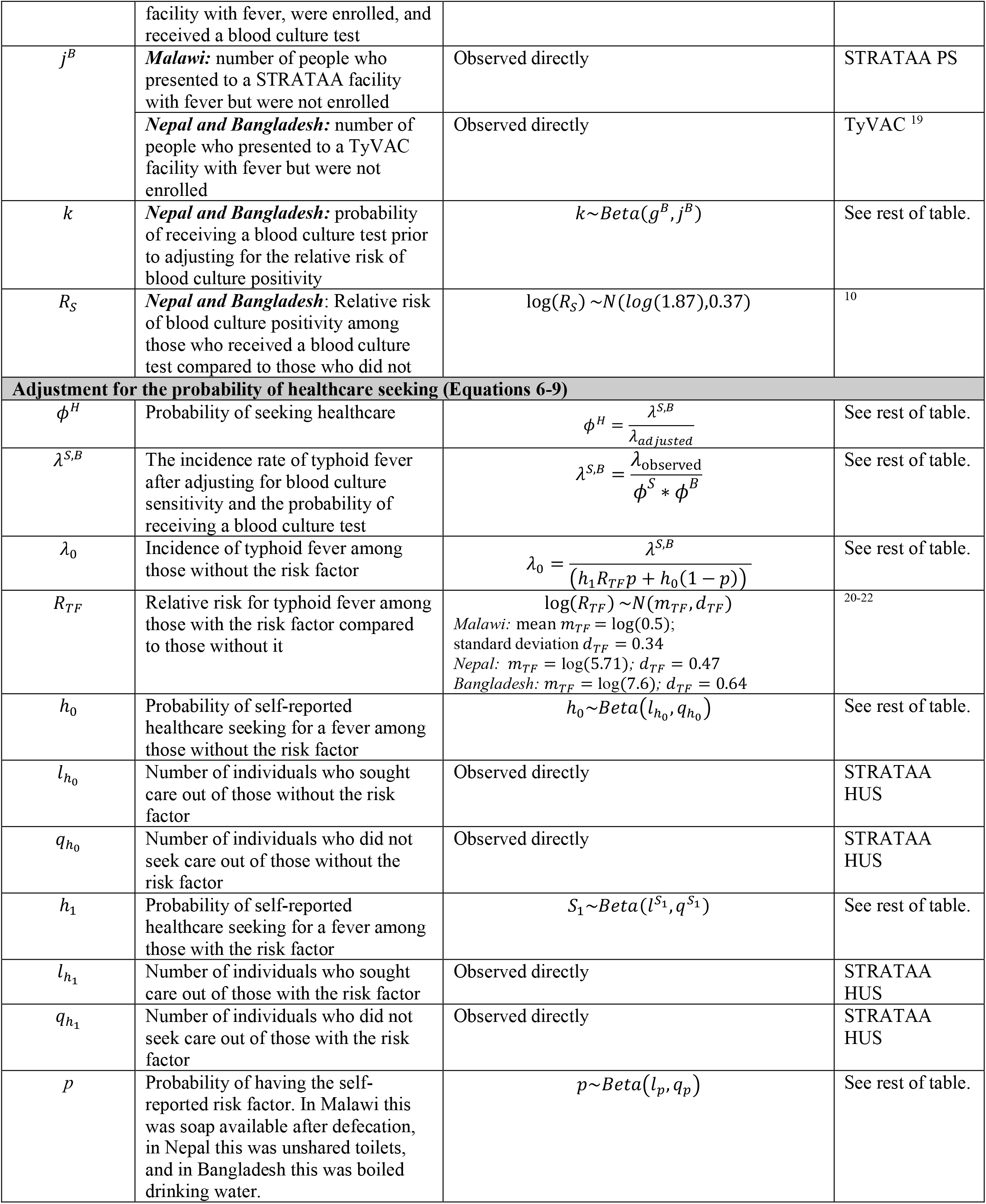
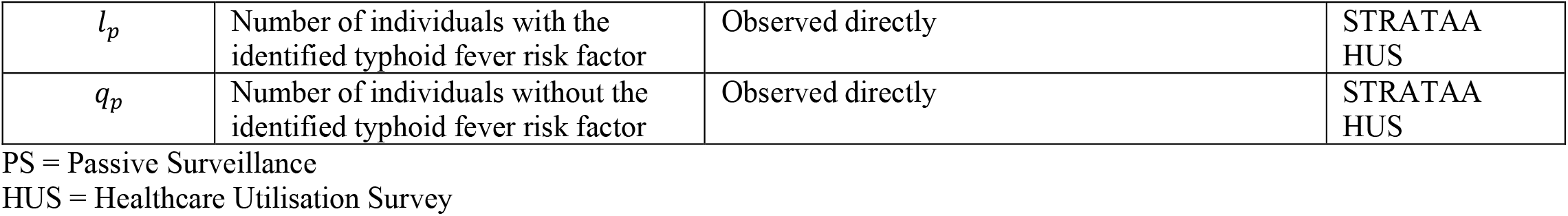
Model input parameters. Parameters used in the incidence adjustment model are described below, with their corresponding uncertainty distributions and data sources. The symbols appear in the table in the order that they appear in the text, organized by the steps in the reporting process (Main model, Adjustment for blood culture sensitivity, Adjustment for the probability of receiving a blood culture test, and Adjustment for healthcare-seeking). All parameters are both age- and country-specific, except for *p* and *R*_*TF*_, for which only a country-level estimate was available. Note that some of parameters in the adjustment for the probability of receiving a blood culture test are specific to only some countries (*ϕ*^*B*^, *g*^*B*^, *j*^*B*^, *k*^*B*^, and *R*_*S*_).

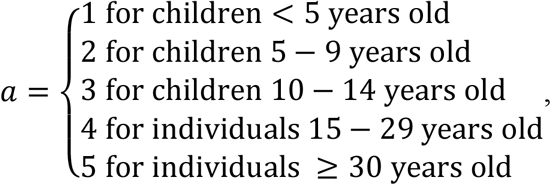

and subscript *c* represents the site, where

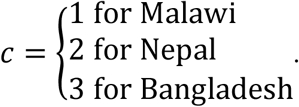

The incidence rate of reported cases can be rewritten as the product of the healthcare-seeking typhoid incidence rate and the probability of a case being captured at each of the three steps of the reporting process,

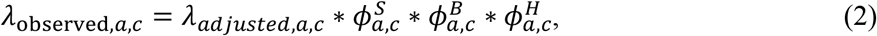

where *λ*_*adjusted,a,c*_ is the final adjusted incidence of typhoid fever incidence, 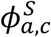 is blood culture sensitivity, 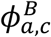 is the probability of receiving a blood culture test, and 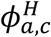 is the probability of seeking healthcare for typhoid fever. Superscript *S* denotes that the parameter refers to the blood culture detection (i.e. sensitivity) phase of reporting, superscript *B* denotes that the parameter is referring to the blood culture collection (i.e. testing) phase of reporting, and superscript *H* denotes that the parameter is referring to the healthcare-seeking phase of reporting.

All model input parameters, with their prior distributions and corresponding data sources, are listed in Table 1. The three steps in the adjustment process (blood culture detection, blood culture collection, and healthcare seeking), are detailed below.

### Adjustment for blood culture sensitivity 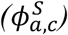

For each individual who received a blood culture test, we inferred whether or not they were a “true” typhoid fever case by adjusting for the specificity and sensitivity of blood culture for typhoid diagnosis (Table S1). First, we assumed that the specificity of blood culture is 100%; thus, all individuals who tested positive for typhoid were assumed to be true cases of the disease. Second, we assumed that among those who tested negative, the probability of being an actual typhoid fever case depended on the volume of blood drawn and prior antimicrobial use, both of which were recorded in the passive surveillance data. Previous studies have shown that the sensitivity of blood culture for typhoid diagnosis is on average 59% (95% CI: 54-64%) but increases by 3% for each additional mL of blood drawn, and decreases by 34% with antibiotic use in the past two weeks ^12^. Thus, each individual who tested negative for typhoid had a probability 1-*w*_*i,v*(*i*),*u*(*i*)_ of being a false negative and thus a true case of typhoid fever, where *w*_*i,v*(*i*),*u*(*i*)_ is the blood culture sensitivity for individual *i, v*(*i*) is the volume of blood collected from individual *i*, and *u*(*i*)=1 if individual *i* reported prior antibiotic use in the previous two weeks and *u*(*i*)=0 otherwise. Blood culture sensitivity is defined as

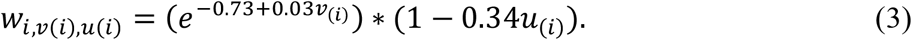

The use or non-use of antibiotics created a bimodal distribution for the sensitivity based on the observed individual-level data calculated using Equation 3 (Fig S2). Thus, we chose to model the distribution of sensitivity among the full population using a normal mixture model with a separate mean and standard deviation for the distribution of blood culture sensitivity with and without the use of antibiotics, varied over blood culture volume. This mixture model is denoted

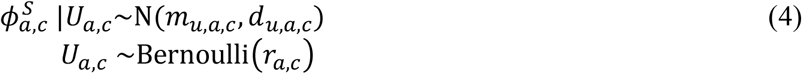

where 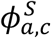 is the blood culture sensitivity adjustment from Equation 2, *ra,c* is the proportion of individuals who took antibiotics in the past two weeks, and *m*_*u,a,c*_ and *d*_*u,a,c*_ represent the mean and standard deviation of the distribution of blood culture sensitivity after adjusting for blood culture volume among those who did (*U*=1) and did not (*U*=0) take antibiotics (from the distribution created using Equation 3), again estimated separately for each age category and site.

### Adjustment for the probability of receiving a blood culture test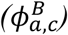

The probability of receiving a blood culture test was estimated differently for Malawi versus Nepal and Bangladesh due to data availability and differences in the primary reasons why individuals were not tested. In Malawi, the main reason why individuals meeting the fever criteria for enrollment did not receive a blood culture test was due to limited capacity and long waiting times at the primary health facility. Individuals often arrived at the clinic early in the morning for clinical review. Once they had seen the government clinician, they were referred for study enrollment and blood-culture collection. If there was a delay in enrollment activities, individuals often left prior to blood cultures being collected. Thus, we assumed that data for those who did not receive a blood culture test were missing completely at random. We used the passive surveillance screening data to estimate the probability of receiving a blood culture test in Malawi, and assumed that the probability followed a beta distribution,

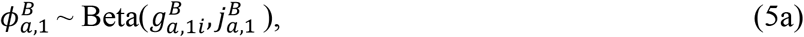

where 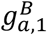 was the number of eligible patients enrolled and 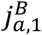 was the number of eligible patients who were not enrolled.

In Nepal and Bangladesh, the primary reason febrile individuals did not receive a blood culture test likely depended on factors associated with their probability of testing positive (e.g., age, number of days of fever, and clinical suspicion of the disease); furthermore, screening data for the passive surveillance were not available. Instead, we relied on published estimates of the relative risk of typhoid fever among those who did not have blood taken for culture and screening data from the Typhoid Vaccine Acceleration Consortium (TyVAC) ^19^. As part of TyVAC, typhoid conjugate vaccine trials are being conducted in the same populations as STRATAA utilizing the same passive surveillance facilities. Baseline information on eligible patients presenting to fever surveillance facilities was recorded both for those who did and did not have blood drawn for culture. Based on the analysis of these data in the TyVAC population in Nepal, the relative risk of blood culture positivity (*R*_*S*_) was 1.87 times higher (95% CI: 0.9-3.9) among those who received a blood culture test compared to those who did not ^10^. Thus, the overall probability of receiving a blood culture test, after taking into account variations in the risk of typhoid fever among those who are and are not tested, is

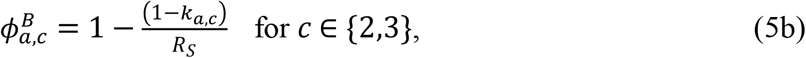

where *k*_*a,c*_ is the probability of receiving a blood culture test prior to adjusting for the relative risk of blood culture positivity. Prior to adjusting for the relative risk, the probability of receiving a blood culture test was assumed to follow a beta distribution, 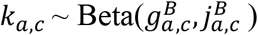, where 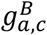 was the number of eligible patients in age category *a* in country *c* who were enrolled and 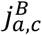 was the number of eligible patients who were not enrolled during TyVAC (for *c* = 2 or 3). While the previous analysis focused only on Nepal, we used the same adjustment for the relative risk of blood culture positivity in Bangladesh, since the reasons for having or not having blood drawn were similar.

### Adjustment for healthcare-seeking behavior 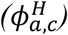

Previous multiplier methods assume that reported healthcare-seeking for a fever is the same as that for typhoid fever; however, this is not necessarily the case. Individuals with typhoid fever may be more or less likely to seek healthcare. In preliminary analyses, we found no difference in reported healthcare seeking by severity of a person’s reported fever, but other factors may explain differential healthcare seeking among those with typhoid fever versus fever due to other causes. To correct for this difference, we measured the probability of seeking care for a fever adjusted for a specified typhoid risk factor to estimate the probability of seeking care for typhoid fever (Fig S1), as described below.

For this phase of reporting, we assumed that everyone in the population either had or did not have a typhoid risk factor, identified from the literature. For each site, we used a different risk factor, based on studies carried out in that specific site and variables for which data was collected as part of STRATAA. The risk factor identified in Malawi was soap available after defecation ^20^; in Nepal, it was unshared toilets ^21^, and in Bangladesh it was boiled drinking water _22_.

In Malawi, the odds ratio for having typhoid fever was 2.0 (95% CI: 1.3-2.5) comparing those who did not use soap after defecation to those who did ^20^. In Nepal, the odds ratio for having typhoid fever was 5.7 (2.3-14.4) comparing those who used a household latrine to those who shared a community latrine ^21^. In Bangladesh, the odds ratio for having typhoid was 7.6 (2.2-26.5) comparing those who did not boil drinking water to those who did ^22^. Since the overall prevalence of typhoid in the population is low, we used these odds ratio estimates from the literature to approximate relative risks for typhoid. We used the same relative risk estimates for all age groups. All other values for the healthcare-seeking adjustment were estimated separately by age and site, as noted below.

We can calculate the marginal probability of seeking care for a fever among those with typhoid fever (alternatively, the incidence of typhoid after adjusting for blood culture sensitivity and the probability of receiving a blood culture test) as:

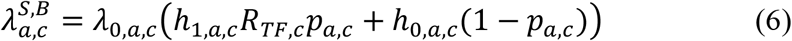

where *p*_*a,c*_ is the probability of having the risk factor for typhoid fever among individuals in age group *a* in site *c, λ*_0,*a,c*_ is the incidence of typhoid fever among those without the risk factor, *R*_*TF,c*_ is the relative risk for typhoid among those with the risk factor (and hence *R*_*TF,c*_*λ*_0,*a,c*_ is the incidence among those with the risk factor), and *h*_1,*a,c*_ and *h*_0,*a,c*_ are the probability of self-reported healthcare seeking for a fever among those with and without the risk factor, respectively.

We can also estimate the overall adjusted incidence of typhoid as the weighted average of the population-based typhoid fever incidence (after adjusting for blood culture sensitivity, the probability of receiving a blood culture test, and healthcare-seeking) among those with or without the typhoid fever risk factor:

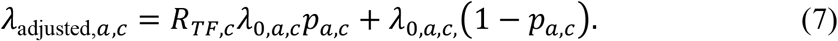

Combining Equations 6 and 7, we can estimate the standardized risk of seeking healthcare for typhoid fever 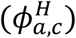 as:

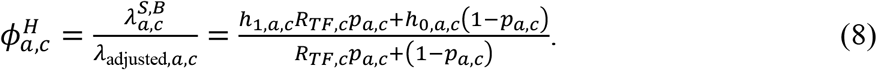

The numbers of individuals with the risk factor and who sought healthcare for a fever were observed directly in a sample of the population in the HUS. We assumed that the probabilities for the occurrence of these numbers (*p, h*_0_, *h*_1_) followed an underlying beta distribution based on the observed values (Table 1).

### Model validation and sensitivity analyses

The final adjusted incidence of symptomatic typhoid fever should be less than or equal to the seroincidence of typhoid infections captured in the serosurveillance data. In this study, seroconversion to typhoid was defined as a ≥ 2-fold rise in anti-Vi IgG titre between the first and second sample drawn and an absolute titre ≥50 EU/ml at the second time point. The seroincidence was estimated as the quotient of the number of people who seroconverted between the first and second blood sample and person-time in years (the number of people sampled multiplied by the mean time between serological samples in age group *a* in country *c*). The final adjusted incidence should fall below the estimated seroincidence rates.

To ensure the model was correctly formulated, we simulated data with known probabilities and incidence rates and compared model estimates to the true values used to generate the data. We simulated high and low values for all of the parameters estimated in the model, starting with a “true” typhoid fever incidence of 1,000 infections per 100,000 pyo. We began by assuming there were 1,250 cases of typhoid fever among 25,000 individuals with a typhoid fever risk factor (*X*_*i*_=1) and 750 cases among 75,000 individuals without the risk factor (*X*_*i*_=0) over two years of surveillance, such that *R*_*TF*_=5. We then sampled from independent Bernoulli random variables, 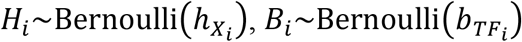, and *U*_*i*_∼Bernoulli(*u*), for the probability of seeking healthcare (***H***), the probability of having blood drawn for culturing (***B***), and the probability of antibiotic usage (***U***). We considered high and low values for each probability, which also varied depending on the risk-factor and typhoid fever status of individual *i* for *h* and *b*, respectively:

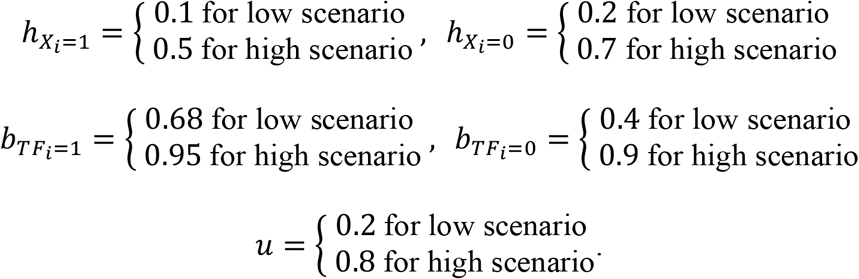

Among individuals with *TF*_*i*_=1 and antibiotic usage *U*_*i*_, the probability of testing positive for typhoid fever (i.e., test sensitivity) was *S*_*i*_∼Bernoulli 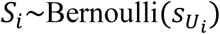, where

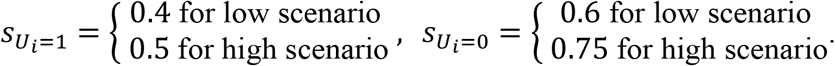

Under both scenarios, the mean test sensitivity was ∼55%. We assumed perfect test specificity, such that *s*_*i*_=0 if *TF*_*i*_=0. The incidence of fever due to other causes was assumed to be 5% per year; we did not allow for multiple episodes of fever over the two-year period, such that

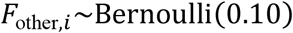

Moreover, we assumed that the incidence of fever due to any cause was

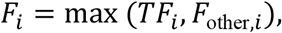

i.e. all individuals could have at most one episode of fever, either due to typhoid fever or another cause. We then generated vectors for whether or not individuals who sought healthcare (***HC***), had blood drawn for culturing (***BC***), and tested positive for typhoid fever (***Y***) as:

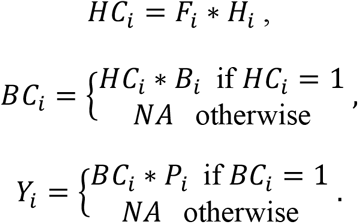

We simulated four scenarios using these probabilities: 1) low probability of seeking care, high probability of being tested, and low prior antibiotic usage; 2) low probability of seeking care, high probability of being tested, and high antibiotic usage; 3) high probability of seeking care, low probability of being tested, and low prior antibiotic usage; and 4) high probability of seeking care, low probability of being tested, and high prior antibiotic usage.

To evaluate whether the final estimates were sensitive to the number of individuals sampled in the HUS, we compared estimates for models that sampled approximately the same number of individuals in each age category as the HUS (735) to models that sampled more individuals (1,000 and 2,000 individuals).

We additionally compared the adjusted incidence estimates from our model to those from a simpler approach that assumed there was no variation in blood culture sensitivity due to prior antibiotic use (i.e., we used a normal distribution for 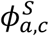 instead of a normal mixture model) and no variation in typhoid incidence among those who were or were not tested (i.e. using the simpler approach to estimate 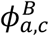 given in Equation 5a) and those who did or did not seek care (i.e. assuming 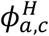 is equal to the probability of seeking care for fever). This simpler approach is more commonly used in multiplier methods for adjustments; however, this approach often faces criticism since it inherently assumes that data are missing completely at random and that reported healthcare-seeking behavior for a fever is the same as that for typhoid fever.

### Model fitting

To estimate the posterior distributions of the adjusted incidence rates, we collected 100,000 posterior samples from the adjustment factors described above following a burn-in period of 10,000 iterations prior to convergence. Convergence was assessed using the Gelman-Rubin diagnostic ^23^ for individual parameters. To ensure the model validation was done without knowledge of the true values, one person simulated the data and another person fit the model to the simulated data. Code for generating simulated data was written in MATLAB version 9.3.0 ^24^. All other analyses were performed using JAGS version 4.3.0 ^25^ in R version 3.4.0 ^26^. Model code, including code for generating the simulated data, is provided at https://github.com/mailephillips/adjusted-typhoid-incidence ^27^.

## Results

The magnitude of the adjustment factors used to estimate the incidence of typhoid fever varied among the three sites (Table 2). In Nepal and Bangladesh, the probability of seeking healthcare was low (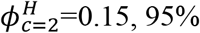credible interval (CrI): 0.09-0.22; and 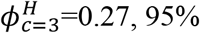 CrI: 0.22-0.33, for all ages) and thus contributed the most to the adjustments, while the probability of receiving a blood culture test when eligible was high (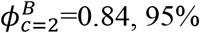 CrI: 0.67-0.92; and 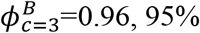 CrI: 0.92-0.98) and contributed the least to adjustments. However, in Malawi, the probability of seeking healthcare was relatively high (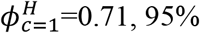 CrI: 0.64-0.77), while the probability of receiving a blood culture test was low (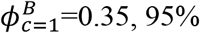 CrI: 0.34-0.36). Blood culture sensitivity was fairly consistent across sites, with median estimates of 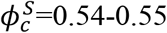.

**Table 2.**
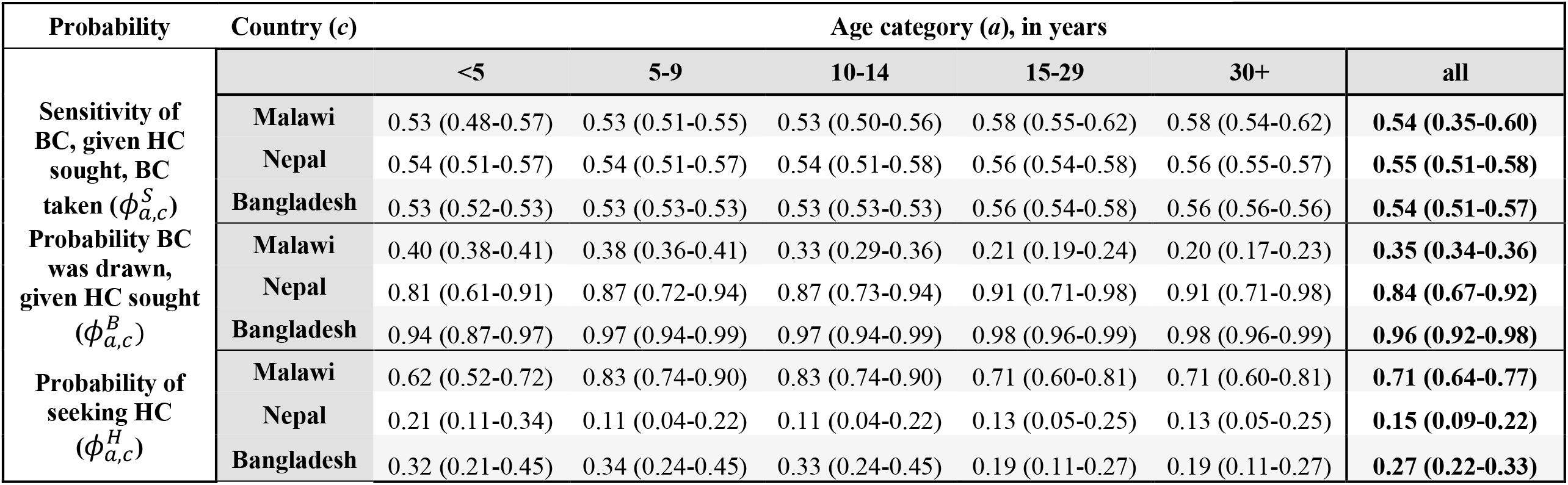
Posterior probability estimates for each adjustment factor by age and site. Each estimate (posterior mean) is shown with its 95% credible interval for the sensitivity of the blood culture (BC) given that healthcare (HC) was sought and a blood culture was taken 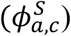, the probability of receiving a blood culture test given that healthcare was sought 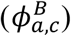, and the probability of seeking healthcare 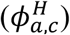. Estimated adjustment factors are shown by age category (*a*) and country (*c*).

The different adjustment factors also varied by age. Blood culture sensitivity was slightly higher in older age groups compared to younger age groups (median estimates of individuals >14 years, 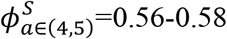; compared to children ≤14 years, 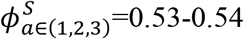). While there was no consistent pattern in prior antibiotic use by age, the amount of blood drawn for a blood culture test generally increased with age. As a result, blood culture sensitivity slightly increased with age. Had we not adjusted for blood culture volume or prior antibiotic use, the estimate for blood culture sensitivity would have been higher for all ages and countries (Fig S5, in blue).

The probability of receiving a blood culture test had different patterns across age groups depending on the country. In Malawi, the probability of receiving a blood culture test decreased with age (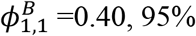 CrI: 0.38-0.41 for children <5 years versus 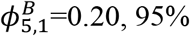 CrI: 0.17-0.23 for adults 30+ years), while in Nepal and Bangladesh, the probability increased (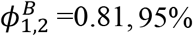 CrI: 0.61-0.91 and 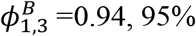 CrI: 0.87-0.97 for children <5 years versus 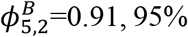 CrI: 0.71-0.98 and 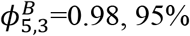 CrI: 0.96-0.99 for adults 30+ years). If we had used a simpler approach (not adjusting for the variation in the risk of typhoid fever among those who were and were not tested) to adjust for the probability of receiving a blood culture test in Nepal and Bangladesh, we would have underestimated the probabilities and thus overestimated the contribution of this adjustment (Fig S5). In Nepal in particular, the simpler approach would have substantially underestimated the probability of receiving a blood culture test among younger age groups. In Bangladesh, the unadjusted proportion of individuals receiving a blood culture test was already close to one, so the adjusted value did not increase the estimate considerably.

The probability of seeking healthcare did not have a consistent pattern across age groups, likely due to the different components contributing to the final estimate. Malawi overall had the highest rates of healthcare seeking, with the lowest rates among children under 5 and the highest rates among children 5-14 (Table 2). Nepal had the lowest rates of healthcare seeking overall, with slightly higher rates among children under 5. Bangladesh also had low healthcare seeking rates, with the lowest rates among adults 15+ years of age (Table 2). In Malawi and Bangladesh, the proportion of those with the relevant typhoid risk factor did not differ by age group (Table S2). Healthcare seeking for a fever was higher among those with the typhoid risk factor in Malawi, but lower among those with the risk factor in Bangladesh and Nepal. When compared to the simpler approach to estimate the probability of healthcare seeking (using the unadjusted proportion of those who sought care for fever), the estimates were similar but slightly higher in most age groups in Malawi but slightly lower in Nepal and Bangladesh (Fig S5).

The magnitude of the overall adjustment to estimate typhoid fever incidence 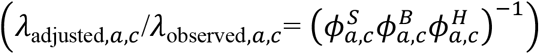 varied between countries and age groups. Nepal had the highest adjustment factors in every age group, with an overall adjustment factor of 14.4 (95% CrI: 9.3-24.9). Malawi and Bangladesh were similar, with adjustment factors of 7.7 (95% CrI: 6.0-12.4) and 7.0 (95% CrI: 5.6-9.2), respectively (Table 3). The highest adjustment factor was for the 5-9-year age group in Nepal (19.7, 95% CrI: 9.0-54.9), while the lowest was for the 5-9- and 10-14-year age groups in Bangladesh (5.8, 95% CrI: 4.1-8.6; and 5.8, 95% CrI: 3.9-8.9, respectively).

**Table 3.** Estimated adjustment factors from final models. The ratio of the median estimate (95% credible interval) of adjusted-to-observed incidence rates is shown for each country and age category.

| Age (years) | Malawi | Nepal | Bangladesh |
| --- | --- | --- | --- |
| **0-4** | 7.6 (4.8-11.6) | 10.7 (4.3-26.8) | 6.3 (4.2-10.2) |
| **5-9** | 5.9 (4.1-8.3) | 19.7 (9.0-54.9) | 5.8 (4.1-8.6) |
| **10-14** | 6.9 (4.3-10.4) | 19.6 (8.6-55.2) | 5.8 (3.9-8.9) |
| **15-29** | 11.4 (6.9-18.0) | 15.8 (7.4-42.4) | 9.8 (6.2-16.7) |
| **30+** | 12.0 (6.0-21.7) | 15.0 (4.6-48.8) | 9.7 (5.5-17.9) |
| **All ages** | 7.7 (6.0-12.4) | 14.4 (9.3-24.9) | 7.0 (5.6-9.2) |

Most of the final adjusted incidence estimates fell within or below the range of the estimated seroincidence of typhoid infection (Table 4). In Malawi, all of the upper bounds of the estimates were well below the seroincidence values. However, in Nepal and Bangladesh, the bounds of the adjusted incidence rates overlapped with the estimated seroincidence. In Nepal, the adjusted incidence among children 5-9 years of age was higher than the seroincidence; however, the confidence/credible intervals overlapped. Similarly, children 10-14 years of age in Bangladesh had higher adjusted incidence rates than seroincidence (which was the lowest across all age groups and sites), but again the confidence/credible intervals overlapped.

**Table 4.** Adjusted typhoid incidence estimates compared to seroincidence. The final adjusted typhoid incidence estimates from the models are shown with 95% credible intervals, as well as the seroincidence estimates with their 95% confidence intervals, by age and site.

| Age | Malawi |  |  | Nepal |  |  | Bangladesh |  |  |
| --- | --- | --- | --- | --- | --- | --- | --- | --- | --- |
|  | Crude rates | Adjusted rates | Seroincidence | Crude rates | Adjusted rates | Seroincidence | Crude rates | Adjusted rates | Seroincidence |
| **0-4 years** | 83<br>(53-124) | 632<br>(398-965) | 2,868<br>(1,153-5,911) | 72<br>(33-136) | 764<br>(307-1,921) | 7,813<br>(2,537-18,232) | 417<br>(337-511) | 2,625<br>(1,764-4,244) | 3,401<br>(1,904-5,610) |
| **5-9 years** | 146<br>(103-201) | 861<br>(599-1,203) | 1,205<br>(146-4,352) | 341<br>(250-455) | 6,713<br>(3,085-18,730) | 5,217<br>(1,915-11,356) | 554<br>(456-666) | 3,228<br>(2,276-4,757) | 3,435<br>(1,571-6,521) |
| **10-14 years** | 88<br>(56-132) | 602<br>(377-915) | 3,061<br>(631-8,946) | 191<br>(128-275) | 3,750<br>(1,653-10,559) | 8,910<br>(4,075-16,916) | 268<br>(203-348) | 1,564<br>(1,050-2,384) | 599<br>(15-3,336) |
| **15-29 years** | 32<br>(20-48) | 361<br>(219-567) | 3,774<br>(1,384-8,213) | 92<br>(71-119) | 1,457<br>(684-3,918) | 10,169<br>(5,255-17,764) | 98<br>(76-124) | 956<br>(603-1,635) | 5,310<br>(2,744-9,275) |
| **30+ years** | 21<br>(10-37) | 248<br>(124-447) | 2,076<br>(762-4,518) | 6<br>(2-13) | 92<br>(29,301) | 7,322<br>(5,100-10,183) | 29 (19-42) | 279<br>(157-514) | 2,988<br>(1,672-4,928) |
| **All ages** | 58<br>(48-70) | 444<br>(347-717) | 2,505<br>(1,605-3,728) | 74<br>(62-87) | 1,062<br>(683-1,839) | 7,631<br>(5,914-9,691) | 161<br>(145-179) | 1,135<br>(898-1,480) | 3,256<br>(2,432-4,270) |

When we evaluated the model against simulated data, the full model was able to reliably estimate both the “true” incidence of typhoid fever and the probabilities used to generate the simulated data for a range of values, while the simpler approach over- or under-estimated the true incidence in some scenarios (Figs 2 and S3). Estimates of blood culture sensitivity were similar to the true value, but incorporated additional uncertainty compared to the simpler approach, consistent with the different sensitivity of blood culture in those who reported prior antibiotic use compared to those that did not (Fig S3). The probability of receiving a blood culture test contributed most to the difference in accuracy between the two approaches. In every scenario, the full model reliably estimated the true value, while the simpler approach underestimated the true value (Fig S3). The adjustment for healthcare seeking in the full model again consistently captured the true value across levels of the probability as compared to the simpler approach, which generally had a narrower 95% CrI that did not always contain the true value (Fig S3). As expected, in both models, the uncertainty in the probability of seeking healthcare decreased as the sampling fraction for the hypothetical HUS increased. As a result, the 95% CrIs in the overall incidence estimates also narrowed as the sampling fraction increased (Fig 2).

**Figure 2.**
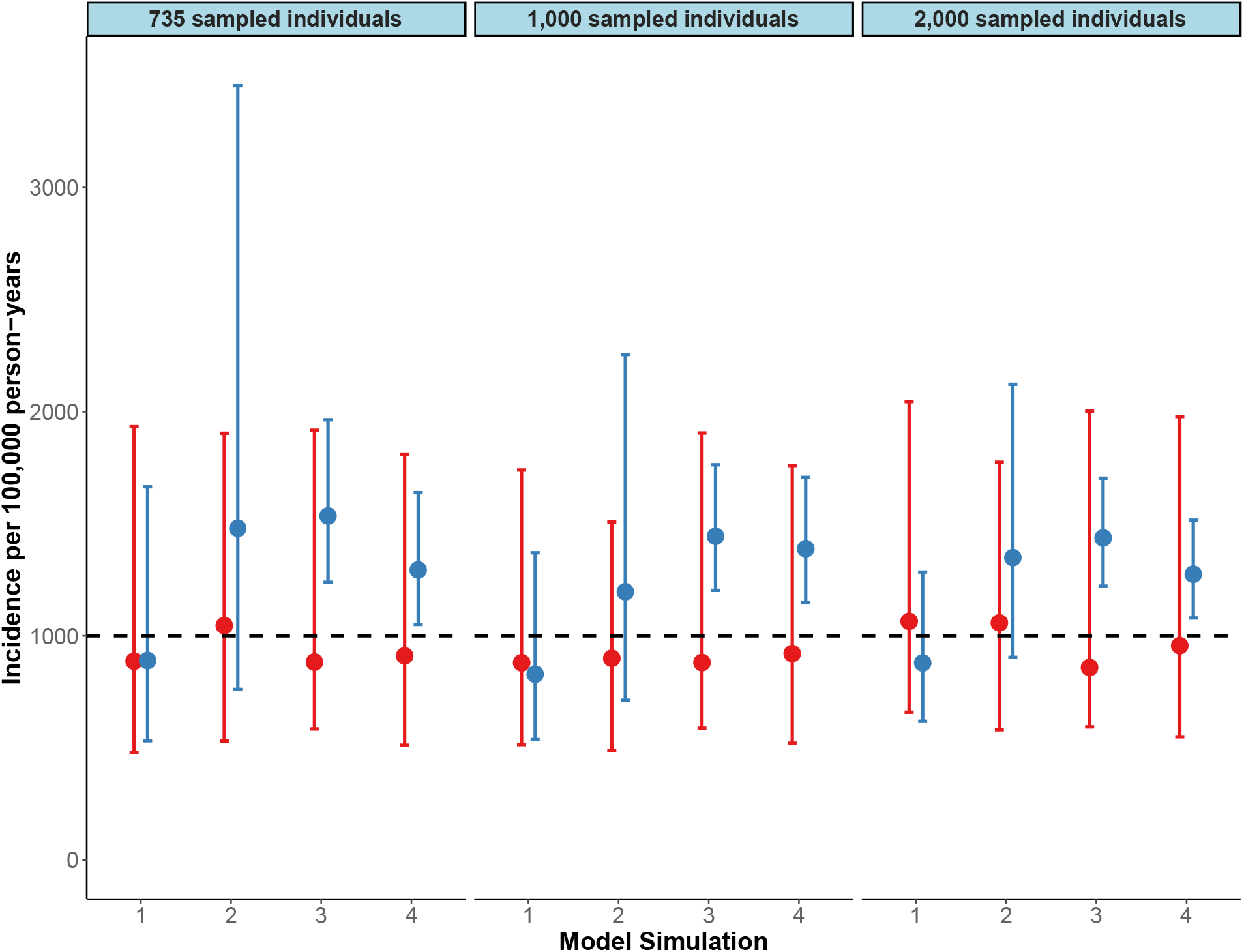
Estimated typhoid incidence based on simulated data: Full model vs. simplified approach. The typhoid incidence per 100,000 person-years of observation was estimated from simulated data based on a true incidence of 1,000 typhoid infections per 100,000 person-years (dashed horizontal black line). Data were simulated for low and high probabilities of seeking healthcare, receiving a blood culture diagnostic test, and antibiotic use. Scenarios were as follows: 1) low probability of seeking care, high probability of being tested, and low prior antibiotic usage; 2) low probability of seeking care, high probability of being tested, and high prior antibiotic usage; 3) high probability of seeking care, high probability of being tested, and low prior antibiotic usage; and 4) high probability of seeking care, high probability of being tested, and high prior antibiotic usage. Each simulation was performed sampling 735; 1,000; and 2,000 individuals from the population for the hypothetical healthcare utilization survey. Estimated “true” values are shown for models that did (red) and did not (blue) account for variation in blood culture sensitivity and variation in typhoid incidence among those who did or did not seek care and were or were not tested.

## Discussion

In order to make informed decisions regarding typhoid control and prevention, it is important to have accurate estimates of population-based typhoid fever incidence. Unreliable reports, inconsistent healthcare utilization, inconsistent clinical diagnoses, suboptimal diagnostic tests, and scarcity of accurate or full data contribute to difficulties in calculating the population-based incidence of typhoid fever. We developed a new methodology within the Bayesian setting to estimate population-based incidence in a context where cases often go undetected and under-reported. Through this approach, we were able to calculate the adjustment factors that can be applied to estimate the “true” incidence of typhoid fever in the STRATAA surveillance sites. These estimates suggested that the adjusted incidence of typhoid fever in Malawi, Nepal, and Bangladesh is 7-to 14-fold higher than the reported blood-culture-confirmed numbers.

It is commonly accepted that cases of typhoid fever go unrecognized at each phase of the reporting process, but the degree to which each step contributes to the underestimation of and uncertainty in the population-based incidence is often not fully quantified. Each of the three intervening processes contributed differently to the underestimation in each country and age group. The probability of seeking healthcare was responsible for the largest portion of underestimation in Nepal and Bangladesh, while the probability of receiving a blood culture test was the biggest factor in Malawi. These results reflect differences in the healthcare systems and fever surveillance processes at the different sites. In Nepal and Bangladesh, antibiotics are widely available and individuals tend to seek care first at a pharmacy instead of a healthcare facility ^28^. In Nepal, a considerable proportion of people with fever neither seek healthcare nor visit a pharmacy possibly due to lack of funds. In Malawi, healthcare seeking is high, but the resources at healthcare facilities are limited. As a result, many people report to healthcare facilities with a fever, receive antibiotics, but do not remain in the facility long enough to receive an additional blood culture test due to prolonged wait times.

Other studies use different approaches to estimate the true incidence of typhoid fever. In many cases, studies simply double the reported cases to account for blood culture sensitivity ^29^. Numerous studies make use of a simple multiplier method ^30-33^, which often do not accurately reflect the uncertainty associated with the reporting process. Previous studies have not attempted to integrate data sources to account for potential differences between the observed healthcare seeking and testing probabilities for fever versus the corresponding unobserved probabilities specific to typhoid fever, which our analysis suggests can impact the final adjustment factors. Our approach can be used to estimate typhoid fever incidence in other study populations. Moreover, some of the issues we encountered (e.g. under-detection due to poor test sensitivity that varies depending patient characteristics, preferential testing of individuals more likely to have the disease of interest, and potential differences in healthcare seeking for those who have the syndrome versus disease of interest) are common to other diseases as well.

By utilizing a Bayesian approach, we were able to measure the contribution to underestimation at each phase of the reporting process while also properly quantifying the uncertainty for each of our estimates. When comparing our model to simulated data, we showed that having more data available (due to higher probabilities of seeking care and receiving a blood culture test) reduced uncertainty in the estimates. Furthermore, we showed that if more people had been sampled in the HUS, uncertainty would also have been reduced.

Our analysis and approach had some limitations. We assumed that healthcare-seeking behavior for fever in households without children is the same as households with children, because the HUS only sampled households with children. Less than a third of households did not have children and were not included in this survey across sites. Studies suggest that households with children are more likely to seek healthcare ^34^, which means that if our estimates are biased, the adjusted incidence estimates are likely conservative. We also were not able to differentiate between febrile illnesses at different parts of the reporting process. We addressed this issue by adjusting for possible differences in healthcare seeking among those with typhoid fever compared to other febrile etiologies using weighted averages based on known risk factors for typhoid fever, but other factors may also lead to differences in healthcare seeking for fever versus for typhoid fever. By comparing our adjusted incidence estimates to estimates of seroincidence, we are able to provide some assurance that the adjusted incidence is within the range of plausible values. However, methods and immunological markers for estimating the seroincidence of typhoid fever are not well established, and the cut-off we used (a ≥2-fold and absolute value of ≥50 EU/mL in anti-Vi IgG) may not be a reliable indicator of acute typhoid infection across all individuals and immunological backgrounds. Another limitation of the approach is that it can very labor-intensive and time-consuming to collect the necessary data.

Calculation of incidence based on data from passive surveillance of blood-culture-confirmed typhoid fever results in considerable underestimation of the true incidence of typhoid fever in the population. Our model provides an approach for estimating typhoid fever incidence while accounting for different sources of information from the reporting process. Typhoid fever remains a major cause of morbidity and mortality in developing countries, so control and prevention are needed. To effectively prioritize, implement and evaluate interventions, estimates of the number of cases should accurately reflect the uncertainty in the reporting process. This analysis provides a platform that can be updated with new or additional data as they become available and can be adapted to other contexts. This model framework could also be used to adjust for underreporting in other diseases.

## Data Availability

Due to patient privacy, the data for this analysis cannot be shared publicly. However, the R code used for the model and the simulated data are available on: https://github.com/mailephillips/adjusted-typhoid-incidence.

## Data availability statement

The data that support the findings of this study are available on request from the corresponding author. The data are not publicly available due to privacy or ethical restrictions.

## Supplementary figures and tables

**Fig S1. Typhoid fever pyramid and febrile pyramid**. The typhoid pyramid (green) is nested within the fever pyramid (grey). Some fraction of symptomatic typhoid fever cases and febrile cases seek care (shaded regions, *TF* and *F*, respectively). The average probability of seeking care for fever is measured (*h;* dashed purple line), but this may vary for individuals with typhoid fever versus fever due to other causes. Within the typhoid fever and fever pyramids, individuals may (X=1) or may not (*X*=0) have a risk factor for typhoid fever; the probability of seeking healthcare varies for those with or without the risk factor, and the risk factor is more prevalent among those with typhoid fever. One can observe whether a person has a fever, but not whether they have typhoid fever.

**Fig S2. Plots of prior antibiotic use, blood culture volume, and blood culture sensitivity**. The average proportion of those with antibiotic use in the past two weeks (A) and the average blood culture volume (B) by country and age group is shown in plots A and B, respectively. In plot C, the distribution of overall (across all age groups) blood culture sensitivity after adjusting for prior antibiotic use and blood culture volume drawn is shown.

**Fig S3. Estimated probabilities from simulated data**. Data for the estimated typhoid incidence were simulated for low, medium and high probabilities of seeking healthcare (*ϕ*^*H*^), receiving a blood culture diagnostic test (*ϕ*^*B*^), and blood culture sensitivity (*ϕ*^*S*^) (row panels); and each simulation was performed sampling 735; 1,000; and 2,000 individuals (column panels) from the population to be “observed” from the healthcare utilization portion. The true values used for simulation are shown in dashed horizontal black lines. The value for blood culture sensitivity without adjusting for prior antibiotic use is shown in dotted gray lines. Estimated values are shown for models that did (red) and did not (blue) account for variation in blood culture sensitivity and variation in typhoid incidence among those who did or did not seek care and were or were not tested.

**Fig S4. Estimated STRATAA typhoid incidence with full model versus a simplified approach**. The estimated typhoid incidence per 100,000 person-years of observation is shown for models that did (red) and did not (blue) take into account variation in blood culture sensitivity and variation in typhoid incidence among those who did or did not seek care and were or were not tested for each age group and country. Note that the upper bounds on children 5-9 and 10-14 in Nepal are not shown.

**Fig S5. Estimated STRATAA probabilities from full model versus a simplified approach**. The estimated probabilities of seeking healthcare (*ϕ*^*H*^receiving a blood culture diagnostic test (*ϕ*^*B*^), and blood culture sensitivity (*ϕ*^*S*^) are shown for models that did (red) and did not (blue) take into account variation in blood culture sensitivity and variation in typhoid incidence among those who did or did not seek care and were or were not tested for each age group and country.

**Table S1. Contingency table of an individual’s sensitivity and specificity for blood culture diagnostic test**. In this study, we assumed that all individuals who tested positive for typhoid fever were true cases of typhoid. Among those who tested negative, an individual *i*’s probability of being a true case of typhoid (*w*_*i,v*(*i*),*u*(*i*)_) depended on the volume of blood drawn *v* and his or her reported prior antibiotic use *u*.

**Table S2. Prevalence of typhoid fever risk factor, rates of reported febrile illness, and probability of healthcare seeking from the Healthcare Utilization Surveys**. The prevalence (prev.) and numerator used to calculate the prevalence (*N*) of each factor used to estimate the probability of healthcare seeking by age and country are show in the table below. Values are shown for the probability of having the risk factor for typhoid fever (*p*), and the proportion of those who sought care at a STRATAA partner health facility among those with (*h*_*1*_) and without (*h*_*0*_) the risk factor are shown.

