## Supplemental Material for "A Bayesian approach for estimating typhoid fever incidence from large-scale facility-based passive surveillance data"

**Short title:** A Bayesian approach for estimating typhoid fever incidence

### Supplementary figures and tables.

**Fig S1. Typhoid fever pyramid and febrile pyramid.** The typhoid pyramid (green) is nested within the fever pyramid (grey). Some fraction of symptomatic typhoid fever cases and febrile cases seek care (shaded regions, *TF* and *F*, respectively). The average probability of seeking care for fever is measured (*h;* dashed purple line), but this may vary for individuals with typhoid fever versus fever due to other causes. Within the typhoid fever and fever pyramids, individuals may (X=1) or may not (*X*=0) have a risk factor for typhoid fever; the probability of seeking healthcare varies for those with or without the risk factor, and the risk factor is more prevalent among those with typhoid fever. One can observe whether a person has a fever, but not whether they have typhoid fever.

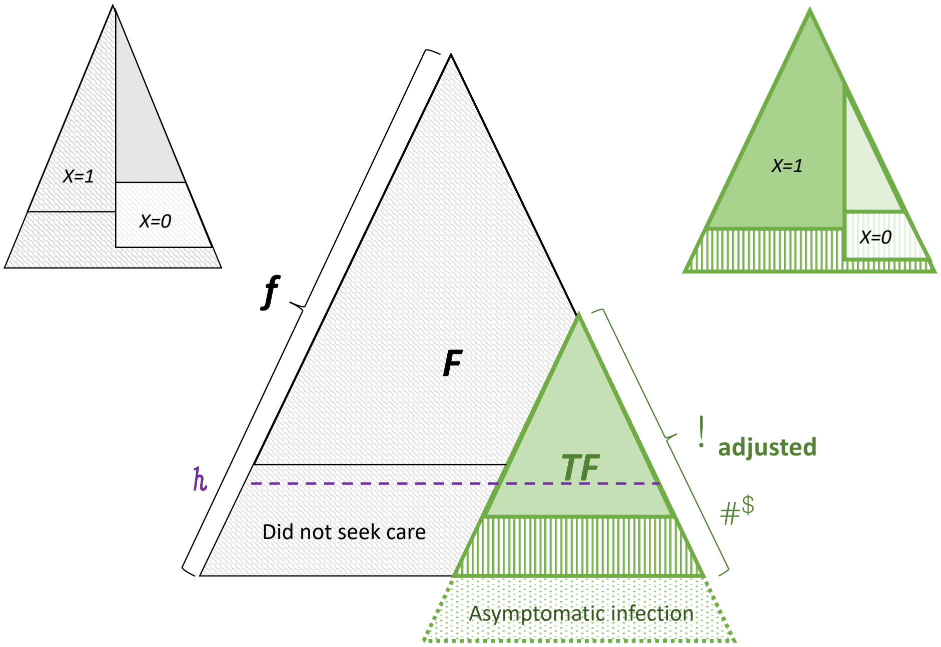

**Fig S2. Plots of prior antibiotic use, blood culture volume, and blood culture sensitivity.** The average proportion of those with antibiotic use in the past two weeks (A) and the average blood culture volume (B) by country and age group is shown in plots A and B, respectively. In plot C, the distribution of overall (across all age groups) blood culture sensitivity after adjusting for prior antibiotic use and blood culture volume drawn is shown.

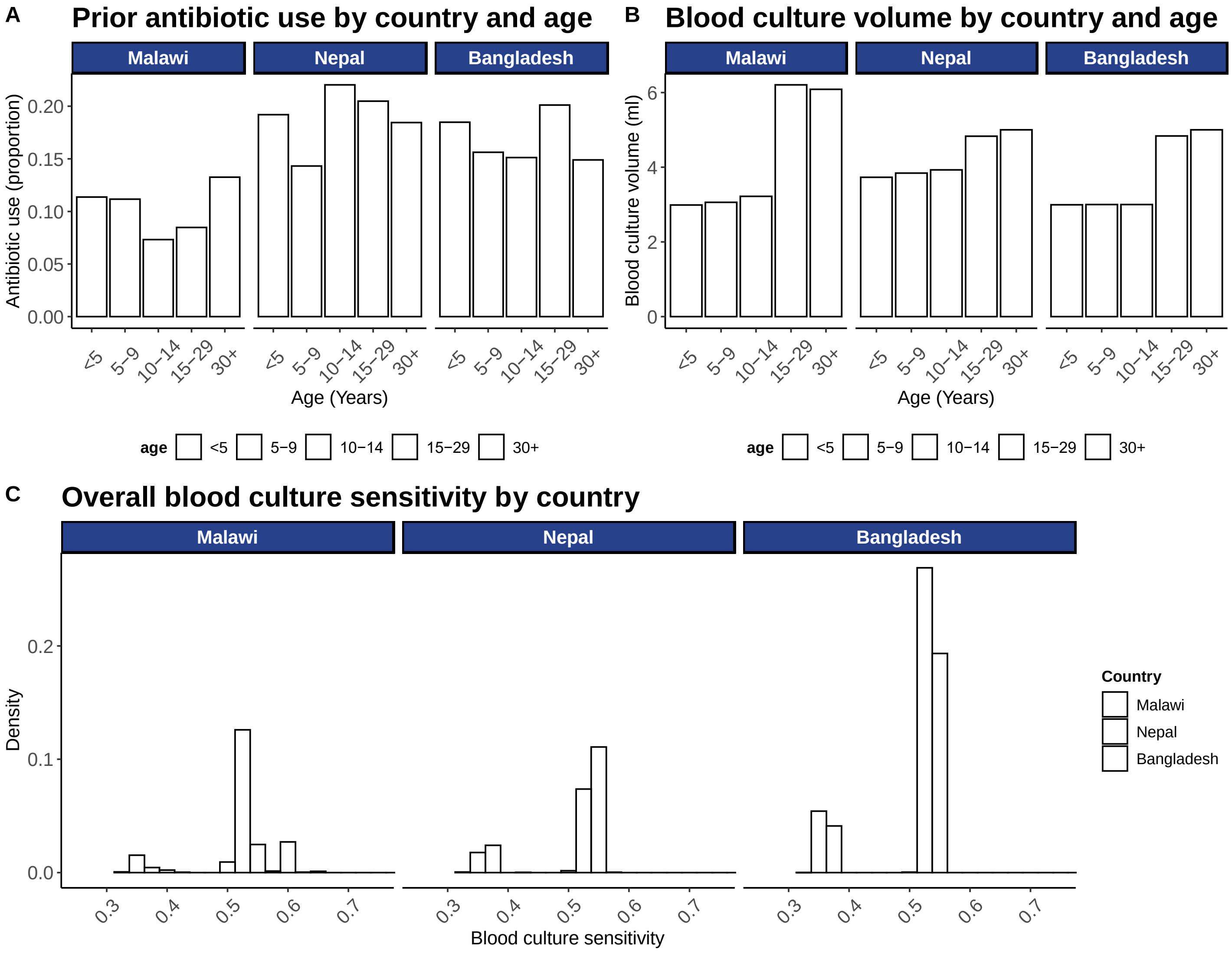

**Fig S3. Estimated probabilities from simulated data.** Data for the estimated typhoid incidence were simulated for low, medium and high probabilities of seeking healthcare ($\phi^{H}$), receiving a blood culture diagnostic test ($\phi^{B}$), and blood culture sensitivity ($\phi^{S}$) (row panels); and each simulation was performed sampling 735; 1,000; and 2,000 individuals (column panels) from the population to be "observed" from the healthcare utilization portion. The true values used for simulation are shown in dashed horizontal black lines. The value for blood culture sensitivity without adjusting for prior antibiotic use is shown in dotted gray lines. Estimated values are shown for models that did (red) and did not (blue) account for variation in blood culture sensitivity and variation in typhoid incidence among those who did or did not seek care and were or were not tested.

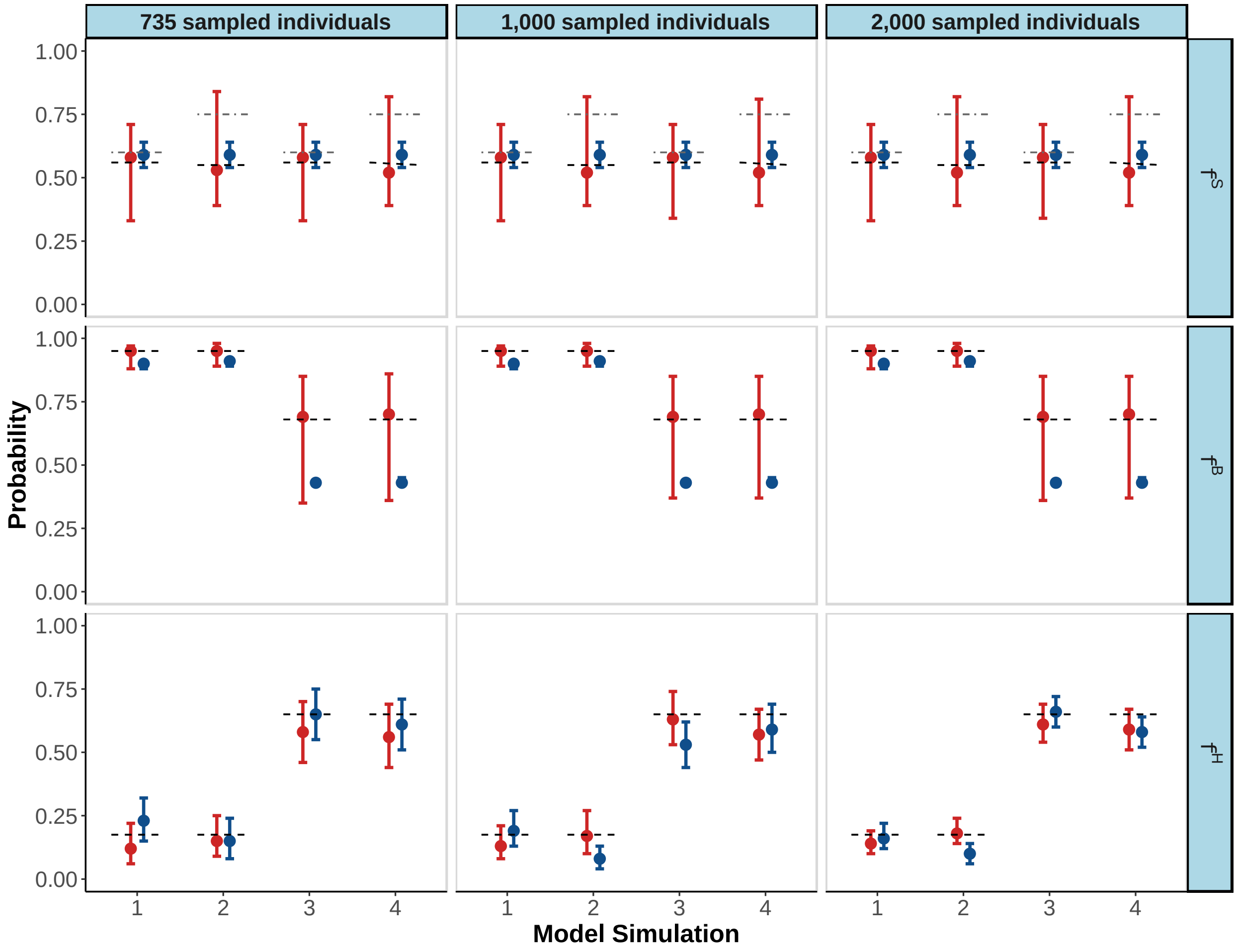

**Fig S4. Estimated STRATAA typhoid incidence with full model versus a simplified approach.** The estimated typhoid incidence per 100,000 person-years of observation is shown for models that did (red) and did not (blue) take into account variation in blood culture sensitivity and variation in typhoid incidence among those who did or did not seek care and were or were not tested for each age group and country. Note that the upper bounds on children 5-9 and 10-14 in Nepal are not shown.

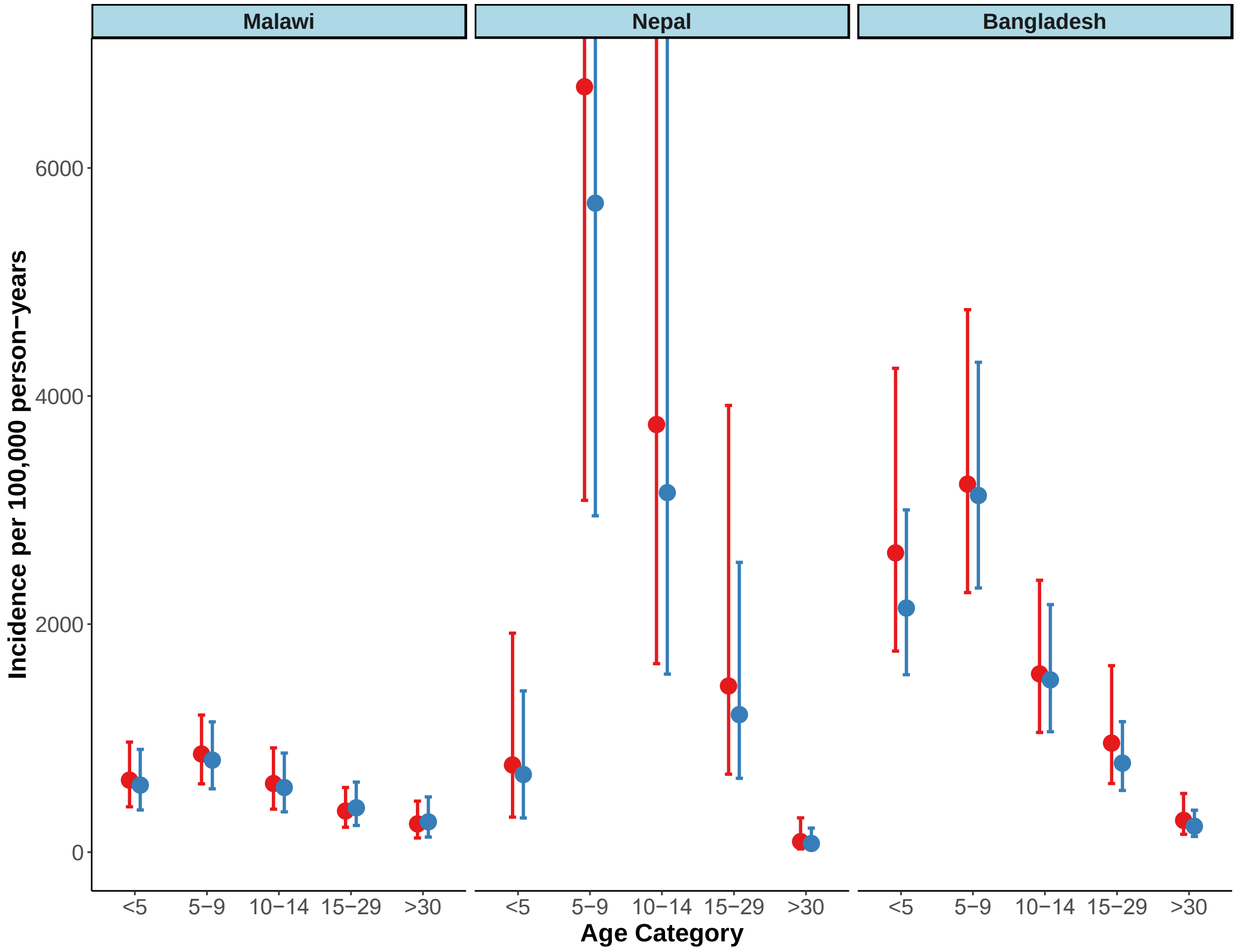

**Fig S5. Estimated STRATAA probabilities from full model versus a simplified approach.** The estimated probabilities of seeking healthcare ($\phi^H$receiving a blood culture diagnostic test ($\phi^B$), and blood culture sensitivity ($\phi^S$) are shown for models that did (red) and did not (blue) take into account variation in blood culture sensitivity and variation in typhoid incidence among those who did or did not seek care and were or were not tested for each age group and country.

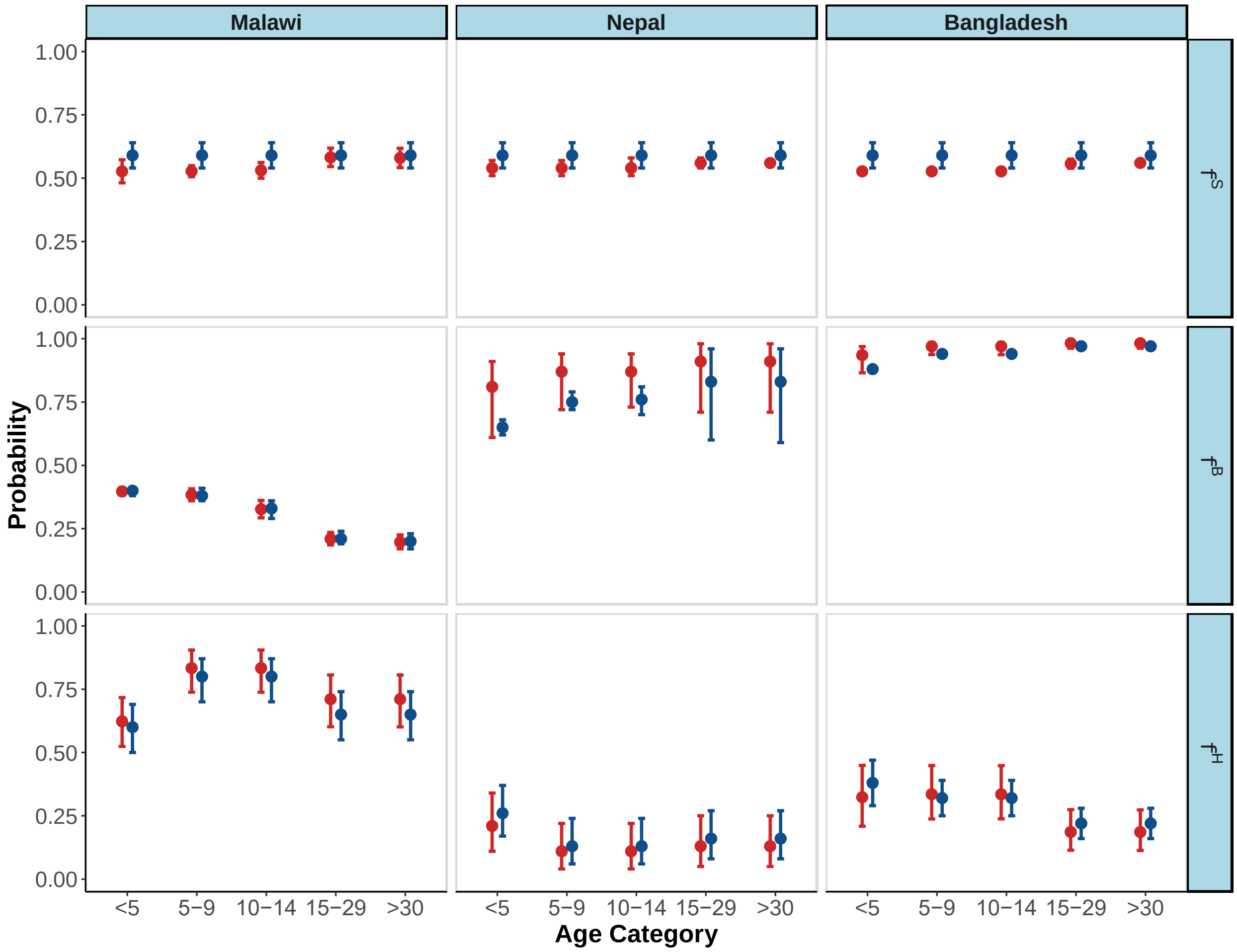

**Table S1. Contingency table of an individual's sensitivity and specificity for blood culture diagnostic test.** In this study, we assumed that all individuals who tested positive for typhoid fever were true cases of typhoid. Among those who tested negative, an individual $i$'s probability of being a true case of typhoid ($w_{i,v(i),u(i)}$) depended on the volume of blood drawn $v$ and his or her reported prior antibiotic use $u$.

| | **Typhoid** | **No typhoid** |
|---|---|---|
| **BC+** | $w_{i,v(i),u(i)}$ | 0 |
| **BC-** | $1 - w_{i,v(i),u(i)}$ | 1 |

**Table S2. Prevalence of typhoid fever risk factor, rates of reported febrile illness, and probability of healthcare seeking from the Healthcare Utilization Surveys.** The prevalence (prev.) and numerator used to calculate the prevalence ($N$) of each factor used to estimate the probability of healthcare seeking by age and country are show in the table below. Values are shown for the probability of having the risk factor for typhoid fever ($p$), and the proportion of those who sought care at a STRATAA partner health facility among those with ($h_1$) and without ($h_0$) the risk factor are shown.

| | | ***p*** | | ***$h_1$*** | | ***$h_0$*** | | ***$R_{TF}$*** |
|---|---|---|---|---|---|---|---|---|
| | ***Age*** | ***prev.*** | ***N*** | ***prev.*** | ***N*** | ***prev.*** | ***N*** | ***Mean (95% CI)*** |
| **Nepal** | ***all ages*** | 0.42 | 130 | 0.13 | 11 | 0.25 | 27 | 5.7 (2.3-14.4) |
| | ***u5*** | 0.37 | 28 | 0.18 | 5 | 0.32 | 15 | |
| | ***5-14*** | 0.45 | 58 | 0.10 | 3 | 0.19 | 5 | |
| | ***15+*** | 0.42 | 44 | 0.12 | 3 | 0.20 | 7 | |
| **Bangladesh** | ***all ages*** | 0.30 | 863 | 0.26 | 35 | 0.31 | 95 | 7.6 (2.2-26.5) |
| | ***u5*** | 0.29 | 146 | 0.29 | 10 | 0.41 | 31 | |
| | ***5-14*** | 0.30 | 306 | 0.35 | 16 | 0.31 | 34 | |
| | ***15+*** | 0.30 | 411 | 0.17 | 9 | 0.24 | 30 | |
| **Malawi** | ***all ages*** | 0.20 | 378 | 0.74 | 59 | 0.64 | 130 | 2.0 (1.3-2.5) |
| | ***u5*** | 0.20 | 126 | 0.72 | 23 | 0.55 | 42 | |
| | ***5-14*** | 0.20 | 126 | 0.78 | 18 | 0.80 | 47 | |
| | ***15+*** | 0.20 | 126 | 0.72 | 18 | 0.62 | 41 | |
